# Diagnostic Accuracy of Simple Postoperative AKI Risk (SPARK) Classification and General Surgery AKI (GS AKI) Index in Predicting Postoperative Acute Kidney Injury among Patients Undergoing Non-Cardiac Surgery at a Tertiary Hospital in the Philippines

**DOI:** 10.1101/2022.03.31.22273255

**Authors:** Michelle Wendy Te, Demi Sarah Robles, Carlo Antonio Boado, Oscar Naidas

## Abstract

**Background:** Postoperative AKI is a significant postoperative complication. Clinical risk prediction models are lacking for patients undergoing non-cardiac surgery. SPARK Classification and GS AKI Index are tools that have shown fair discriminative ability to predict post-operative AKI in non-cardiac surgery and have external validation in their original cohorts. There is no study that compares the diagnostic accuracy of both tools.

**Objectives:** This study aims to compare the diagnostic accuracy of SPARK Classification vs GS-AKI Risk Index in predicting post-operative AKI among patients who will undergo non-cardiac surgery at a tertiary hospital in the Philippines.

**Methods:** This is a cross-sectional study, including adult patients who underwent non-cardiac surgeries from January 2019 to July 2021. The individual risk of post-operative AKI for both models were determined. Descriptive data was described using t-test and logistic regression. Measures of accuracy were described using sensitivity, specificity, positive and negative predictive value, positive and negative likelihood ratio, and discriminative ability using concordance (c) statistic.

**Results:** Of the 340 patients in this study, 77 (22.65%) developed post-operative AKI and 24 (7.06%) developed critical AKI. Based on demographic data, older age, pre-existing renal disease, longer duration of surgery, anemia, hypoalbuminemia, and hyponatremia were associated with higher incidence of post-operative AKI. SPARK had a sensitivity ranging from 17-43% and specificity ranging from 58-93% for Class B to C. GS AKI had a sensitivity ranging from 10-26% and specificity ranging from 61-97% for Class I to V. SPARK had a discriminative power (c statistic) ranging from 0.46 to 0.61 while GS AKI had a discriminative power ranging from 0.41 to 0.54.

**Conclusion:** Based on this study, there is an association between higher risk classification in both SPARK and GS AKI and postoperative AKI. However, both clinical prediction models demonstrate poor discriminative power to predict post-operative AKI.

## Introduction

Postoperative Acute Kidney Injury (AKI) is an undesired complication in surgical patients of variable outcome. Strategies to prevent AKI using practical, externally validated risk prediction indices are integral to both identification of at-risk patients prior to surgery and their subsequent management of post-surgery. Among developed prediction tools, many have been validated to accurately predict post-operative AKI in cardiac surgeries, however these are lacking for non-cardiac surgeries. A common risk prediction tool that we use is the General Surgery AKI Risk Index developed by Kheterpal et al.[1] with good discriminative ability. The SPARK Classification developed by Park et al.[2] is a new risk prediction tool that includes more variables in its model than GS AKI, and further classifies outcomes as critical AKI, which they defined as AKI stage 2 as defined by KDIGO, AKI requiring renal replacement therapy, and AKI leading to death. This risk prediction model has limited generalizability because it was only validated in the Korean population. There is a need to validate this test in the local setting. If proven to be more specific and sensitive than GS-AKI Risk Index, the SPARK Classification could replace GS AKI as the standard tool to predict the risk of postoperative AKI.

## Review of Related Literature

### Epidemiology of AKI in Non-cardiac Surgical Patients

Acute kidney injury (AKI) is characterized by a sudden decrease in renal function from injury to the kidneys. The diagnosis of AKI is based on the Kidney Disease Improving Global Outcomes (KDIGO) criteria defined by increase in serum creatinine or decrease in urine output.[3]

Acute kidney injury is a common complication associated morbidity and risk for mortality. AKI has diverse etiologies and early detection and diagnosis is paramount to early treatment and reversal. The estimated global burden of AKI was 31% in Southeast Asia and the pooled mortality of AKI was 13.8%[4]. This indicates that the numbers reveal an enormous medical burden in the region.

AKI was found to be prevalent in both general hospital and intensive care unit settings. In the region, male predominance is observed at 55-75% and was associated with chronic comorbid conditions such as hypertension, diabetes and chronic kidney disease.[4]

The most common cause of AKI is sepsis and closely in second is surgery. Postoperative-AKI (PO-AKI) is defined as AKI within seven days of surgery. AKI occurring more than seven days after surgery are not necessarily related to the surgery itself hence should be managed as hospital-acquired AKI.[5]

The incidence of AKI in patients undergoing cardiac surgery is estimated to be at 42% depending on the complexity of the surgery and use of bypass.[6] In cases of abdominal surgery, rates are noted to be up to 13% and the risk is noted to be increased depending on additional patient-related risk factors for development of AKI like chronic vascular disease, chronic kidney disease, arterial hypertension, cardiac failure, and diabetes.[7] In comparison, AKI is seen in after orthopedic (6.7%), thoracic (12.1%), vascular surgery(9.3%), and urologic surgeries (8.6%).[8]

Among the causes of AKI, the leading cause of in-hospital AKI was renal hypoperfusion and nephrotoxic agents. In Southeast Asia, community acquired AKI contributes more to the burden of illness. The exact cause of CA-AKI was not studied in the SEA but among neighboring countries, the most common cause were infectious diseases (including leptospirosis) and sepsis.

In a study involving 110 Filipino patients aiming to describe the clinical profile and risk factors for mortality of patients with acute renal failure, the overall mortality rate was 14% with most deaths occurring on the first week of confinement. AKI in the study was associated with older age, male gender, and the presence of comorbid illness. The most common cause of AKI was noted to be infections such as leptospirosis and decreased renal perfusion.[9]

Risk assessment is an essential step to mobilize interventional strategies to minimize further renal injury. The non-recognition rate of AKI ranges has a high rate resulting in delay in nephrology referral and was shown to increase mortality rates, HD dependence and hospital stay.

### Outcomes of AKI in Surgical Patients

A number of studies showed that AKI had negative impact on outcomes of surgical patients which included increased mortality, healthcare cost, longer hospital stay and increased likelihood of dialysis dependence. Several studies showed that increase in creatinine alone was not the sole cause of poor outcomes in surgical patients with AKI. Among surgical patients, the systemic multifactorial pathophysiology causing organ dysfunction is central in postoperative AKI. [10] Those who develop AKI may have consequences after hospitalization, with greater risk of developing CKD and higher incidence of long-term mortality. Chronic inflammation, transformation of pericytes into myofibroblasts in response to injury, and build-up of extracellular matrix leading to permanent scarring and changes in renal function.[11]

### Pathophysiology and Diagnosis of AKI in Non-cardiac Surgery Patients

Perioperative AKI is multifactorial and is commonly precipitated by several insults which most commonly involves hypoperfusion and inflammation. Hypovolemia during surgery may develop at intra-operatively or post-operatively which reduced the mean arterial pressure reducing renal blood flow. Persistent hypotension results into a decrease in glomerular filtration rate secondary to afferent and efferent vasoconstriction above the effects of ADH and Angiotensin II. Systemic inflammatory reaction from the surgery results into tubular dysfunction and injury causing AKI. Tubular damage is caused by microcirculatory dysfunction, leukocyte migration, and endothelial dysfunction.[12]

### Risk Factors to Development of AKI

A multitude of risk factors were identified to contribute to the development of AKI in patients undergoing non-cardiac surgery. These risk factors are divided into patient- and surgical-related factors. Table 1 describes the multiple risk factors for postoperative AKI.

**Table 1.** Perioperative Risk Factors for Postoperative AKI (Modified from Molinari, L 2020)

| Pre-operative Factors | Intraoperative Factors | Post operative Factors |
| --- | --- | --- |
| Preoperative<br>Preoperative hospital stay > 5d<br>Preoperative sepsis<br>Race<br>Male sex<br>Age<br>High ASA Status<br>High RCRI<br>Comorbidities<br>CKD<br>DMT2<br>COPD<br>Smoking<br>CHF and Hypertension<br>Arterial and venous vascular disease<br>CAD<br>Prior MI<br>Arterial fibrillation<br>Liver disease<br>Cancer<br>Bleeding disorders<br>Medications<br>Diuretics<br>NSAIDs<br>Nephrotoxic antibiotics<br>Steroids<br>Contrast agents<br>ACEi/ARBs<br>Pre-operative laboratory<br>eGFR reduction<br>Anemia<br>Hypoalbuminemia<br>High CRP<br>Acidosis HCO<sub>3</sub> < 22 | Emergent surgery<br>Laparotomy<br>Abdominal surgery<br>IAH<br>Longer duration of surgery<br>IV Contrast<br>Hemodynamic instability<br>Use of diuretics<br>Transfusion<br>Invasive Procedures<br>Vascular surgery<br>No use of cold renal perfusion<br>Open surgery<br>Anatomic complexity<br>Bleeding > 1L<br>Hemoglobin < 10 g/dL<br>Transplant surgery<br>Cirrhosis<br>CKD<br>Immunosuppression | Sepsis<br>Hemodynamic instability<br>Low cardiac output<br>Hypotension<br>Hypovolemia<br>Respiratory failure<br>Mechanical ventilation<br>Leak of anastomosis<br>Positive fluid balance within 48h<br>Transfusion<br>Use of diuretics<br>Use of vasopressor<br>Use of NSAIDs<br>Resumption of ACE 15 days post surgery |

Chronic diseases like CKD, chronic obstructive pulmonary disease, and diabetes are well established risk factors in the development of AKI. Other factors include obesity which increases the odds to develop AKI by 26.5% per 5 kg/m2 BMI. Use of nephrotoxic agents like antibiotics, non-steroidal anti-inflammatory medications also increase the risk. Other comorbidities are important because these patients may have creatinine values within the normal range but decreased GFRs.[6] In these patients, it is conceivable that less serious insults might have more detrimental effects as in patients without comorbidities. Hemodynamic alterations such as prolonged periods of hypotension and vasopressors also increase the risk of AKI. The type of surgery such as general, thoracic, orthopedic, vascular, and urological surgeries are further worth mentioning as an association with the development of AKI has been described.[13]

The detection of patients at risk for AKI plays a pivotal role because these patients might particularly benefit from preventive strategies.

### Use of Clinical Prediction Scores

The use of clinical prediction scores for AKI is a frequent topic amongst nephrologists. There are available validated prediction scores for AKI for patients undergoing cardiac surgery, non-cardiac surgery, and orthopedic surgery. The most established are for cardiac surgery.

According to Lei, V et al.[14], existing clinical prediction rules for postoperative AKI provided only moderate levels of accuracy. These studies differ in their definitions of the AKI outcome, use of a mix of statistical and machine learning approaches, and have not uniformly focused on non-cardiac surgery.

### General Surgery Acute Kidney Injury (GS-AKI) Risk Index

The clinical tool developed by Kheterpal involving 75,952 non-cardiac operations in 2009 was derived from the database of outcomes from general surgery procedures performed in 121 medical centers in the US. The primary outcome of the study included AKI within 30 days, defined as an increase in serum creatinine of at least 2 mg/dl or dialysis requirement.[1] A list of pre-existing comorbidities and operative characteristics were evaluated as possible predictors of AKI. A logistic regression was used to create an AKI model. The study identified five risk factors: age >56 years, male sex, active congestive heart failure, ascites, hypertension, emergency surgery, intraperitoneal surgery, renal insufficiency - mild or moderate, and diabetes mellitus (oral or insulin therapy).

After defining the risk factors for AKI, they categorized the patients according to the number of risk factors each patient has: class 1 (0-2 risk factors), class II (3 risk factors), class III (4 risk factors), class IV (5 risk factors), and class V (6+ risk factors). The incidence of AKI was determined for each class, which is 0.2%, 0.8%, 1.8%, 3.3%, and 8.9%, respectively. The corresponding hazards ratio of each class was determined with their corresponding p-value. The discriminative ability (c statistic) for a simplified risk index was 0.80 in the derivation and validation cohorts. Class V patients (six or more risk factors) had a 9% incidence of AKI. Overall, patients experiencing AKI had an eightfold increase in 30-day mortality.[15]

### Simple Postoperative AKI Risk (SPARK) Classification

Recently, the development of a prediction tool addressed postoperative complications, including AKI across a range of surgical settings. The Simple Postoperative AKI Risk (SPARK) classification is a validated preoperative AKI risk score for non-cardiac surgery developed in South Korea, involving 51,041 patients and validated in 39,764 patients.[2] The classification used nine pre-operative variables: age, sex, baseline GFR, urinary albuminuria, expected surgery duration, emergency operation, diabetes mellitus, renin-angiotensin-aldosterone system blockade usage, hypoalbuminemia, anemia, and hyponatremia.

The SPARK risk score considers AKI severity by predicting a composite of adverse renal outcomes which included: development of stage 2 or greater AKI termed as “Critical AKI”, which was defined as either a need for RRT within 90 days of AKI or death occurring after any AKI diagnosis. The tool was developed to assist clinicians to discuss to patients the risk before surgery in a clear and definite quantification of risk and compilations versus the benefits of the surgery.

Higher risk patients can also be put on intensive monitoring and interventions in the post-operative period. This risk prediction model however has limited generalizability because it was only validated in Korean population hence additional validation is required to test its applicability to other populations. The discriminative power of the SPARK index was acceptable in both the discovery (c-statistic 0.80) and validation (c-statistic 0.72) cohorts.

### Web Based Prediction Model for AKI after Surgery

A recent study developed and validated a risk prediction tool for the occurrence of postoperative AKI requiring RRT (AKI-dialysis). The study involved 2,299,502 surgical patients from the American College of Surgeons National Surgical Quality Improvement Program Database (ACS NSQIP) from 2015 to 2017. The model involved 11 predictors including: age, history of congestive heart failure, diabetes, ascites, emergency surgery, hypertension requiring medication, preoperative serum creatinine, hematocrit, sodium, preoperative sepsis, and surgery type. The risk model was developed using multivariable logistic regression. The AKI risk prediction model had high discriminative power for postoperative AKI-dialysis (c statistic training cohort: 0.89, c statistic test cohort: 0.90). The calculator provides only risk estimate and individual surgical outcome. [16]

### Study Objectives

The aim of this study was to compare the accuracy of SPARK Classification vs GS-AKI Risk Index in predicting post-operative AKI among patients who will undergo noncardiac surgery at St. Luke’s Medical Center - Quezon City.

In order to do this, we aimed to evaluate the accuracy in terms of sensitivity, specificity, predictive values, likelihood ratios, and to determine the ROC (c statistic) of the SPARK Classification and GS-AKI Risk Index in predicting postoperative AKI among patients who will undergo noncardiac surgery at SLMC – QC; to determine if there is a significant difference in the accuracy indices of the SPARK Classification and GS-AKI Risk Index in predicting postoperative AKI among patients who will undergo noncardiac surgery at SLMC – QC; and to present the clinical profile of patients referred for Nephrology risk stratification from year 2019 to 2021.

## Materials and Methods

### Ethical Approval

The Institutional Ethics Review Committee of St. Luke’s Medical Center - Quezon City approved the study (EC Reference number: SL-20133). Informed consent was waived as the study was a cross-sectional study without medical intervention.

### Study Hospital and Study Design

This was a cross-sectional study performed in St. Luke’s Medical Center - Quezon City, which included adult (age ≥ 18 years) patients who underwent non-cardiac surgeries from January 2019 to July 2021. The exclusion criteria were as follows: (1) cardiac surgeries, (2) ophthalmologic and dermatologic surgeries, (3) nephrectomy or kidney transplant recipients, (4) obstetric procedures, (5) minor procedures defined as surgery duration <1 hour, (6) patients who were administered local anesthesia, (7) patients who had established or preoperative kidney dysfunction, defined as patients with CKD stage V or undergoing hemodialysis, and (8) patients with incomplete study variables found in medical record.

### Operational Definitions

### Study Procedure

Patients who underwent non-cardiac surgery were identified from the general surgery census of St. Luke’s Medical Center - Quezon City starting January 2019 to July 2021. The medical records were reviewed for eligibility into this study. The following data were extracted: age, sex, active congestive heart failure, hypertension, diabetes mellitus, use of ACE/ARBs, surgical procedure, expected surgical duration, reported as emergency operation, serum creatinine, hemoglobin, serum albumin, urinalysis, presence of ascites, postoperative serum creatinine, incidence of new onset renal replacement therapy, and death. Charts with incomplete data were excluded. Preoperative data were extracted from the medical records, and both SPARK and GS AKI were applied to every patient. The postoperative outcomes were extracted from the same records.

### Study Outcome

A post-operative AKI event was defined as AKI occurring within 7 days of an operative intervention using the KDIGO definition of AKI (defined as any of the following: (1) increase in serum creatinine by ≥ 0.3mg/dL within 48 hours, (2) increase in serum creatinine to ≥ 1.5 times baseline occurring within 7 days, or (3) urine volume <0.5ml/kg/h for 6 hours). To enhance the clinical use of the study findings, we grouped the outcomes into two categories: low risk to intermediate risk AKI (SPARK Class A and B, GS AKI Class I to IV) and high risk AKI (SPARK Class C and D, GS AKI Class V). Among patients who developed post-operative AKI, critical AKI was defined as AKI stage ≥ 2 and AKI that led to post-AKI death or dialysis within 90 days.

### Study Sampling and Sample Size

Both GS-AKI and SPARK have Area under curve (c discrimination) of 0.80 based on the study of Kheterpal (2009) and Park (2019). Thus in order to compare the two tools, as assumption of a difference of 0.15 in their AUC and ratio of 10 (based on 13.2% incidence of AKI on the study of Cheng), the minimum sample size required is at least 31 patients with AKI and 310 without AKI. This is based on 95% confidence level and 80% power of test.

### Data Analysis

The study demographics and perioperative characteristics were summarized using descriptive statistics. Categorical data was expressed as frequency and proportions, while continuous data was expressed as mean. For bivariate analysis, a two-tailed t test was used to compare the differences between means of two groups. The variables that were identified from SPARK and GS AKI were put into a univariate binary logistic regression model between the dependent and independent variables, and were compared using odds ratio. The accuracy indices for both SPARK and GS AKI were computed and expressed as sensitivity, specificity, positive predictive value, negative predictive value, positive and negative likelihood ratio. The discriminative ability of both models were measured using the concordance statistic (c statistic) and presented under the Receiver Operating Characteristic (ROC) curve.

## Results

The demographic, clinical, and surgical profiles of the respondents according to post-operative AKI status can be found on Table 3. It can be noted that those who developed post-operative AKI had a mean age of 68.14 years old (SD=12.84), while those who did not develop post-operative AKI had a mean age of 58.49 years old (SD=14.89). Comparative analysis indicated that the mean age of those who developed post-operative AKI was significantly higher (t=–5.18, p=0.001). Results also showed that majority of those who developed post-operative AKI were males (53.85%), had hypertension (56.41%), and had an estimated glomerular filtration rate (eGFR) of more than 60mL/min/1.73m2 (66.67%). It is also notable that among those who developed post-operative AKI, 7.69% had congestive heart failure, 33.33% had diabetes mellitus, 26.92% used ACE or ARB medications, and 5.19% had ascites. In contrast, among those who did not develop post-operative AKI, 56.27% were females, 13.69% had pre-existing renal disorders, 80.23% had an eGFR of more than 60mL/min/1.73m2, 5.32% had congestive heart failure, 47.91% had hypertension, 27.00% had diabetes mellitus, 23.57% used ARB or ACE medications, and 5.13% had ascites. Comparative analyses indicated no significant difference in these characteristics between those who developed and did not develop post-operative AKI (p>0.05), except for existing renal diseases (χ2=3.92, p=0.048).

**Table 2.** Nine variables included in the General Surgery AKI Index. [1]

| Risk Factors for GS-AKI |
| --- |
| Age greater than or equal to 56 years old |
| Male sex |
| Active congestive heart failure |
| Ascites |
| Hypertension |
| Emergency Surgery |
| Intraperitoneal Surgery |
| Renal insufficiency – mild or moderate |
| Diabetes Mellitus – oral or insulin therapy |

**Table 3.** Perioperative PO-AKI risk evaluation strategy on the basis of the suggested SPARK classification. [2]

| Preoperative Risk Factors | Scores |
| --- | --- |
| Age (years) |  |
| < 40 | 0 |
| > or equal to 40 and <60 | 6 |
| > or equal to 60 and <80 | 9 |
| > or equal to 80 | 13 |
| eGFR (ml/Min per 1.73m<sup>2</sup>) |  |
| > or equal to 60 | 0 |
| > or equal to 45 and <60 | 8 |
| > or equal to 30 and <34 | 15 |
| > or equal to 15 and < 30 | 22 |
| Dipstick albuminuria (urine albumin +1) | 6 |
| Sex |  |
| Female | 0 |
| Male | 8 |
| Expected surgical duration (hours) | x 5 |
| Emergency operation | 7 |
| Diabetes Mellitus | 4 |
| RAAS Blockade | 6 |
| Hypoalbuminemia (< 3.5g/dL) | 8 |
| Anemia (<12g/dL females, <13g/dL males) | 4 |
| Hyponatremia (<135mEq/L) | 3 |

**Table 4.** Operational definitions.

|  | Definition |
| --- | --- |
| Age | 56 years and older |
| Sex | Male or female |
| Active congestive heart failure | Abnormal exercise tolerance, orthopnea, increased JVP, Paroxysmal nocturnal dyspnea, pulmonary rales on physical examination, cardiomegaly, pulmonary vascular engorgement Noted in the medical record as congestive heart failure or pulmonary edema |
| Ascites | The presence of fluid accumulation in the peritoneal cavity noted on physical examination, abdominal ultrasound, or abdominal computed tomography or magnetic resonance imaging within 30 days before the operation |
| Hypertension | Persistent elevation of systolic blood pressure of >140mmHg or a diastolic blood pressure of > 90 mmHg or requires antihypertensive treatment at most within 30 days of being considered as a candidate for surgery |
| Emergency surgery | No later than 12 hours after the patient has been admitted to the hospital or after the onset of related preoperative symptomatology, OR if surgeon or anesthesiologist reported case as emergent |
| Intraperitoneal surgery | Incision reaches the peritoneum, excluding hernia repairs |
| Renal insufficiency - mild or moderate | Mild renal insufficiency: serum creatinine 1.2 to 1.9mg/dL Moderate renal insufficiency: serum creatinine of 2.0 mg/dL or greater |
| Diabetes mellitus - oral or insulin therapy | 2 categories: DM necessitating oral hypoglycemic therapy OR DM necessitating insulin therapy |
| Dipstick albuminuria | Presence or absence of urine albumin greater than or equal to 1+ |
| Hypoalbuminemia | Serum albumin less than 3.5g/dL |
| Anemia | Hemoglobin < 12g/dL in females or <13g/dL for males |
| Hyponatremia | Serum sodium less than 135 mEq/L |

In terms of surgical characteristics, results indicated that the duration of surgery was significantly longer (t=–3.49, p=0.001) among those who developed post-operative 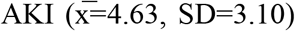 than those who did not 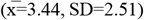. Results also showed that the proportion of patients who had emergency surgery and developed post-operative AKI (2.56%) was not significantly different (χ2=0.41, p=0.740) from those who did not develop post-operative AKI (4.20%).

The pre-operative characteristics of the respondents according to post-operative AKI status is presented in Table 6. It can be noted that the proportion of patients who had pre-operative anemia was significantly higher (χ2=4.19, p=0.027) among those who developed post-operative AKI (70.51%) than those who did not (56.49%). Similarly, the proportion of patients who had pre-operative hypoalbuminemia was significantly higher (χ2=4.39, p=0.036) among those who had post-operative AKI (78.21%). Results also indicated that the proportion of patients who had pre-operative hyponatremia was significantly higher (χ2=4.91, p=0.027) among those who had post-operative AKI (38.46%) than those who did not develop the post-operative outcome (25.19%). Results showed that the proportion of patients with dipstick hypoalbuminemia and the mean pre-operative serum creatinine was not statistically different between those who developed and did not develop post-operative AKI (p>0.05).

**Table 5.** Demographic, Clinical, and Surgical Profiles of the Respondents according to Post-Operative Acute Kidney Injury (AKI) Status (N = 340)

| Characteristics | Post-Operative Acute Kidney Injury (AKI) Status |  |  | Test Statistic | p-value (Two-Tailed) |
| --- | --- | --- | --- | --- | --- |
|  | With Post-Operative AKI (n = 78) | Without Post-Operative AKI (n = 262) | Total (N = 340) |  |  |
| Age ($\bar{x}$, SD) | 68.14 (12.84) | 58.49 (14.89) | 60.73 (14.97) | -5.18<sup>†</sup> | 0.001 |
| Sex (f, %) |  |  |  | 2.56 | 0.108 |
| Female | 36 (46.15%) | 114 (43.51%) | 184 (54.12%) |  |  |
| Male | 42 (53.85%) | 148 (56.49%) | 156 (45.88%) |  |  |
| **Clinical Data** |  |  |  |  |  |
| Pre-Existing Renal Disease (f, %) | 18 (23.08%) | 36 (13.74%) | 54 (15.88%) | 3.92* | 0.048 |
| Estimated Glomerular Filtration Rate (mL/min/1.73m<sup>2</sup>; (f, %) |  |  |  | 9.37* | 0.024 |
| <15 | 0 (0.00%) | 0 (0.00%) | 0 (0.00%) |  |  |
| >15 to <30 | 3 (3.85%) | 11 (4.20%) | 14 (4.12%) |  |  |
| >30 to <45 | 8 (10.26%) | 15 (5.73%) | 23 (6.76%) |  |  |
| >45 to <60 | 15 (19.23%) | 23 (8.78%) | 38 (11.18%) |  |  |
| >60 | 52 (66.67%) | 213 (81.30%) | 264 (77.65%) |  |  |
| Congestive Heart Failure (f, %) | 6 (7.69%) | 14 (5.34%) | 20 (5.88%) | 0.60 | 0.439 |
| Hypertension (f, %) | 44 (56.41%) | 125 (47.71%) | 169 (49.71%) | 1.82 | 0.177 |
| Diabetes Mellitus (f, %) | 26 (33.33%) | 71 (27.10%) | 97 (28.53%) | 1.15 | 0.284 |
| Use of ACE or ARB Medications (f, %) | 21 (26.92%) | 61 (23.28%) | 82 (24.12%) | 0.44 | 0.509 |
| Presence of Ascites (f, %) | 4 (5.13%) | 12 (4.58%) | 16 (4.71%) | 0.04 | 0.768 |
| **Surgical Characteristics** |  |  |  |  |  |
| Duration of Surgery (Hours; $\bar{x}$, SD) | 4.63 (3.10) | 3.44 (2.51) | 3.71 (2.70) | -3.49<sup>†</sup> | 0.001 |
| Emergency Surgical Procedure (f, %) | 2 (2.56%) | 11 (4.20%) | 13 (3.82%) | 0.41 | 0.740 |

\*Significant at 0.05
<sup>†</sup>Significant at 0.01

**Table 6.**
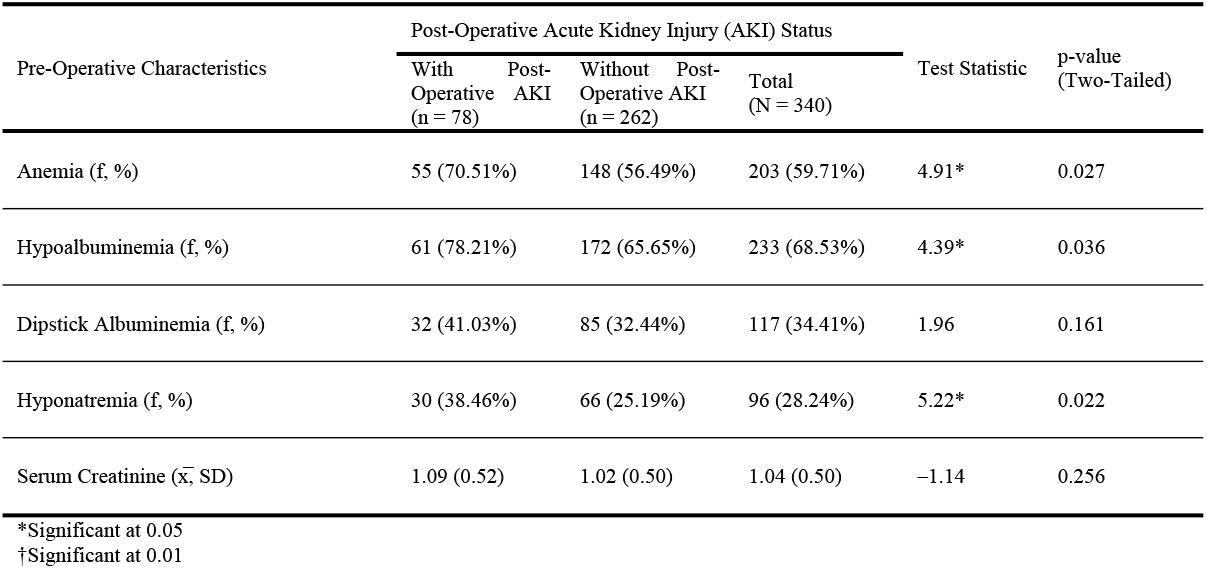
Pre-Operative Characteristics of the Respondents according to Post-Operative Acute Kidney Injury (AKI) Status (N = 340)

The post-operative outcomes of the respondents according to post-operative AKI status is depicted in Table 7. It can be gleaned from the table that the proportion of patients who developed critical AKI (30.77%), needed renal replacement therapy within 90 days (21.79%), and expired (21.79%) was significantly higher (p<0.05) among those who had developed post-operative AKI. Likewise, results showed that the mean post-operative serum creatinine was significantly higher (t=–13.38, p=0.001) among those who had post-operative AKI than those who did not. Further, those who had post-operative AKI showed a significant increase in post-operative creatinine, when compared with pre-operative levels (t=–14.91, p=0.001), with a mean percentage change of 83.88% (SD=85.44%) than those who did not have post-operative AKI 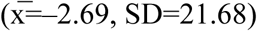.

**Table 7.**
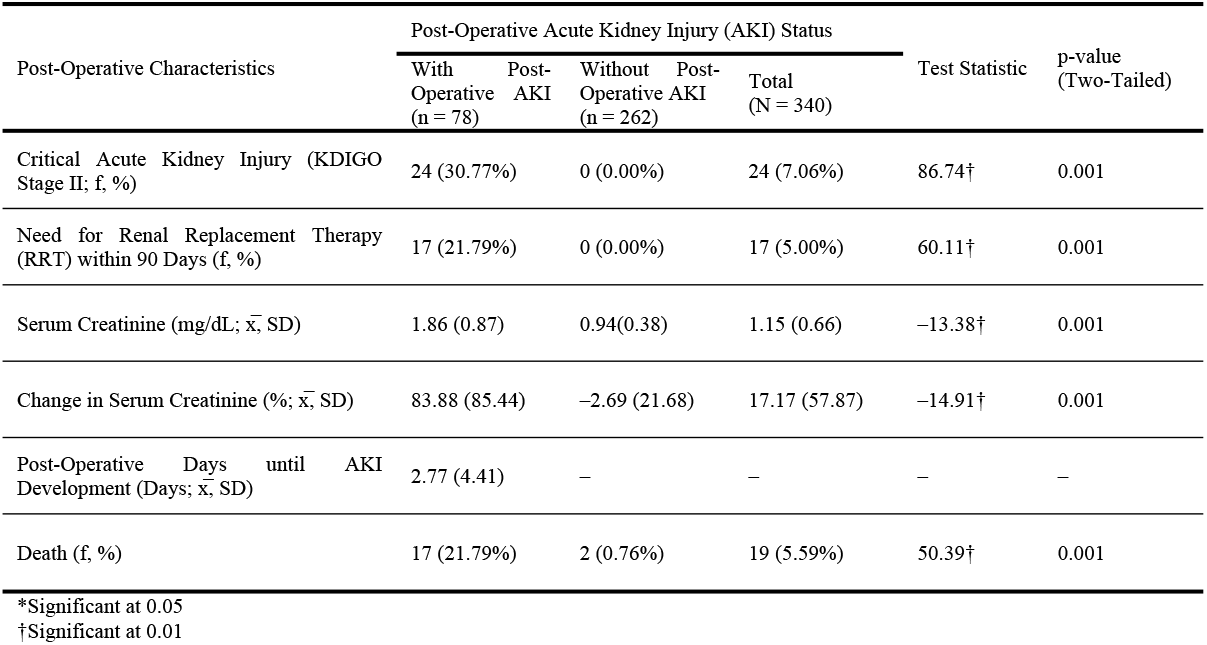
Post-Operative Outcomes of the Respondents according to Post-Operative Acute Kidney Injury (AKI) Status (N = 340)

Table 8 illustrates the AKI risk stratification using the Simple Postoperative AKI Risk (SPARK) Classification and the General Surgery AKI (GS-AKI) Index among the respondents according to post-operative AKI status. Results indicated that among those who developed post-operative AKI, 82.05% were categorized as high risk using the SPARK Classification and 10.26% were classified as high risk using the GS-AKI index. Only 18.18% were of low to intermediate category using the SPARK classification and 89.61% were considered low to intermediate risk using the GS-AKI Index. Comparative analyses indicated that the proportion of patients classified as high risk was significantly higher among those who developed post-operative AKI, in both the SPARK Classification (χ2=14.37, p=0.001) and GS-AKI Index (χ2=8.44, p=0.008).

**Table 8.**
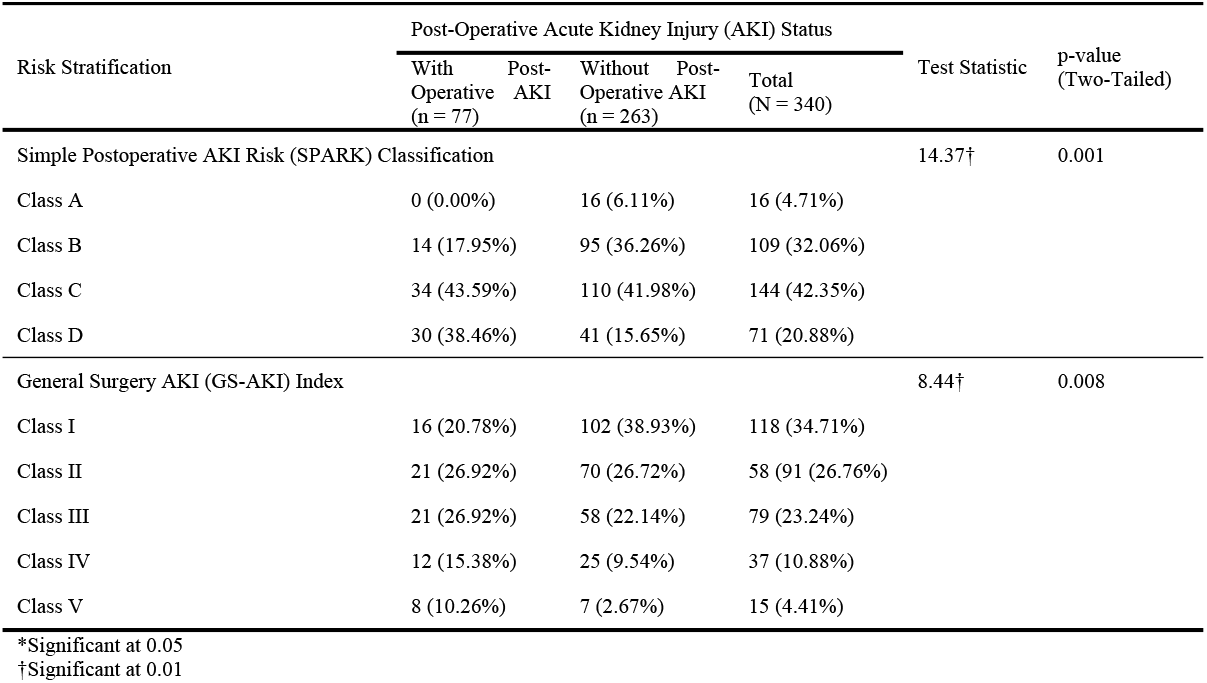
Frequency Distribution of the Acute Kidney Injury (AKI) Risk Stratification using the Simple Postoperative AKI Risk (SPARK) Classification and the General Surgery AKI (GS-AKI) Index among the Respondents according to Post-Operative Acute Kidney Injury (AKI) Status (N = 340)

The diagnostic capacity of the different categories of the Simple Postoperative AKI Risk (SPARK) Classification and the General Surgery AKI (GS-AKI) Index are illustrated in Table 9 and Figure 1. Using receiver operating characteristic (ROC) curve (Figure 4), it can be noted that the AUC-ROC of the different classes of the SPARK Classification ranged from 40.80% to 61.40%, while the different categories of GS-AKI Index had accuracy values, also known as c-statistic, ranging from 40.80% to 53.80%. The AUC-ROC curve, which reflects the accuracy of the screening tool, is conventionally categorized into five: fail (≤0.60), poor (0.61 to 0.70), fair (0.71 to 0.80), good (0.81 to 0.90) and excellent (≥0.91). Using these values, the different classes of the SPARK Classification had a failure or poor diagnostic accuracy, while the different GS-AKI classifications had failed to accurately screen post-operative AKI.

**Table 9.** Diagnostic Properties [Accuracy, Sensitivity, Specificity, Positive Predictive Value (PPV), Negative Predictive Value (NPV), and Likelihood Ratios (LR)] of the Simple Postoperative AKI Risk (SPARK) Classification and the General Surgery AKI (GS-AKI) Index with Post-Operative Acute Kidney Injury (AKI) among the Respondents in Screening Post-Operative Acute Kidney Injury (AKI) among the Respondents (N = 340)

| Risk Assessment<br>Questionnaire<br>Score Categories | Post-Operative Acute Kidney Injury (AKI) |  |  |  |  |  |  |
| --- | --- | --- | --- | --- | --- | --- | --- |
|  | AUC-ROC<br>(95% CI) | Sensitivity<br>(95% CI) | Specificity<br>(95% CI) | PPV<br>(95% CI) | NPV<br>(95% CI) | LR+<br>(95% CI) | LR-<br>(95% CI) |
| **Simple Postoperative AKI Risk (SPARK) Classification** |  |  |  |  |  |  |  |
| Class A | 0.469<br>(0.455 – 0.484) | 0.00%<br>(0.00% – 4.62%) | 93.90%<br>(90.30% – 96.50%) | 0.00%<br>(0.00% – 20.60%) | 75.90%<br>(70.90% – 80.50%) | 0.00<br>(0.00 – 0.00) | 1.07<br>(1.03 – 1.10) |
| Class B | 0.408<br>(0.357 – 0.460) | 17.90%<br>(10.20% – 28.30%) | 63.70%<br>(57.60% – 69.60%) | 12.80%<br>(7.20% – 20.60%) | 72.30%<br>(66.00% – 78.00%) | 0.50<br>(0.30 – 0.82) | 1.29<br>(1.12 – 1.48) |
| Class C | 0.508<br>(0.445 – 0.571) | 43.60%<br>(32.40% – 55.30%) | 58.00<br>(51.80% – 64.10%) | 23.60%<br>(16.90% – 31.40%) | 77.60%<br>(71.10% – 83.20%) | 1.04<br>(0.78 – 1.39) | 0.97<br>(0.78 – 1.21) |
| Class D | 0.614<br>(0.555 – 0.673) | 38.50%<br>(27.70% – 50.20%) | 84.40%<br>(79.40% – 88.50%) | 42.30%<br>(30.60% – 54.60%) | 82.20%<br>(77.00% – 86.50%) | 2.46<br>(1.65 – 3.66) | 0.73<br>(0.61 – 0.88) |
| **General Surgery AKI (GS-AKI) Index** |  |  |  |  |  |  |  |
| Class I | 0.408<br>(0.354 – 0.462) | 20.50%<br>(12.20% – 31.20%) | 61.10%<br>(54.90% – 67.00%) | 13.60%<br>(7.95% – 21.10%) | 72.10%<br>(65.70% – 77.90%) | 0.53<br>(0.33 – 0.84) | 1.30<br>(1.12 – 1.51) |
| Class II | 0.501<br>(0.445 – 0.557) | 26.90%<br>(17.50% – 38.20%) | 73.30%<br>(67.50% – 78.50%) | 23.10%<br>(14.90% – 33.10%) | 77.10%<br>(71.40% – 82.20%) | 1.01<br>(0.66 – 1.53) | 1.00<br>(0.86 – 1.16) |
| Class III | 0.524<br>(0.468 – 0.579) | 26.90%<br>(17.50% – 38.20%) | 77.90%<br>(72.30% – 82.70%) | 26.60%<br>(17.30% – 37.70%) | 78.20%<br>(72.70% – 83.00%) | 1.22<br>(0.79 – 1.87) | 0.94<br>(0.81 – 1.09) |
| Class IV | 0.529<br>(0.485 – 0.573) | 15.40%<br>(8.21% – 25.30%) | 90.50%<br>(86.20% – 93.70%) | 32.40%<br>(18.00% – 49.80%) | 78.20%<br>(73.10% – 82.70%) | 1.61<br>(0.85 – 3.06) | 0.94<br>(0.84 – 1.04) |
| Class V | 0.538<br>(0.503 – 0.573) | 10.30%<br>(4.53% – 19.20%) | 97.30%<br>(94.60% – 98.90%) | 53.30%<br>(26.60% – 78.70%) | 78.50%<br>(73.60% – 82.80%) | 3.84<br>(1.44 – 10.30) | 0.92<br>(0.85 – 1.00) |

\*Significant at 0.05
†Significant at 0.01

**Fig 1.**
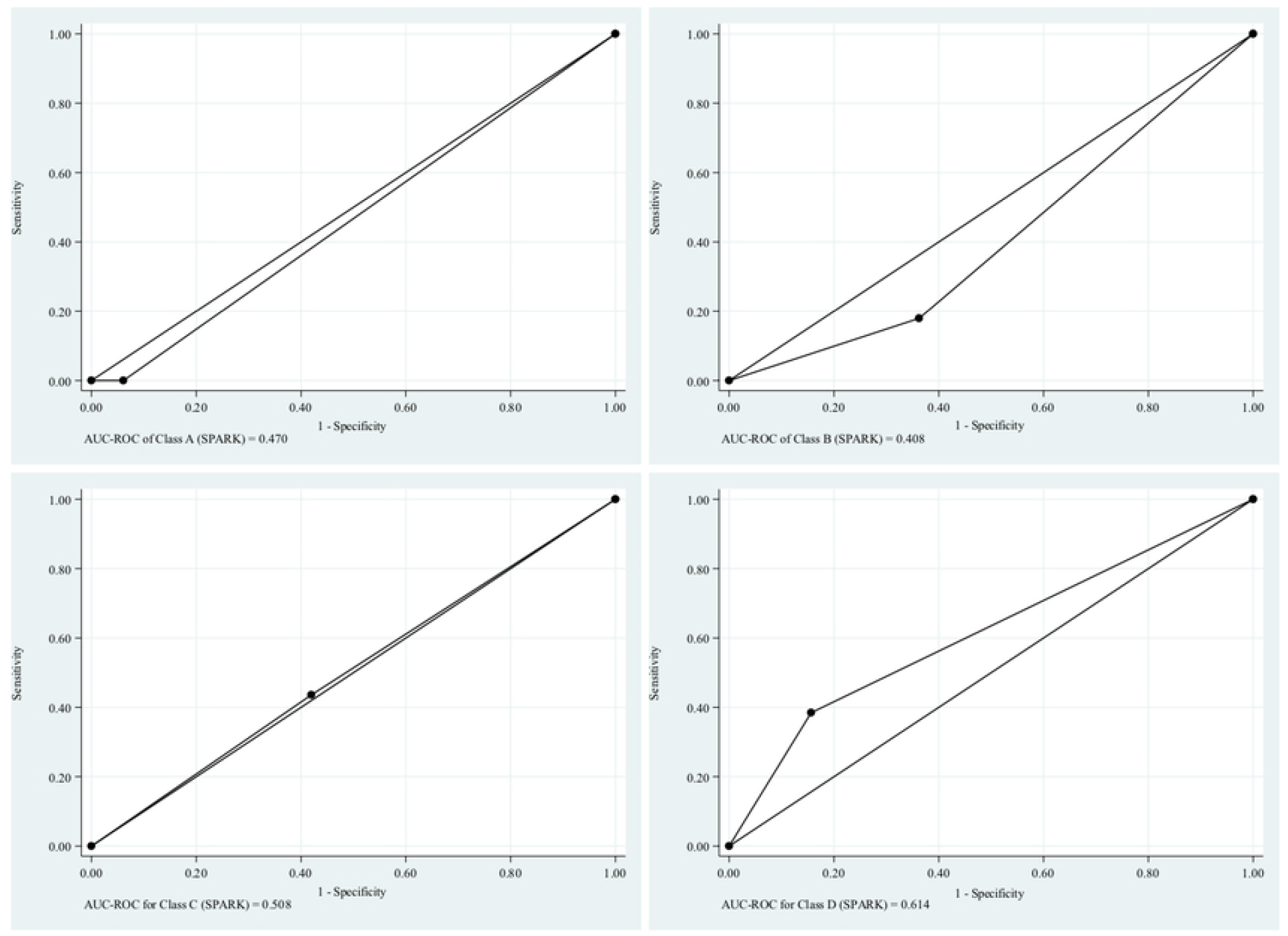
Area Under the Receiver Operating Characteristic (AUC-ROC) of the different GS-AKI Categories (N = 340)

**Fig 2.**
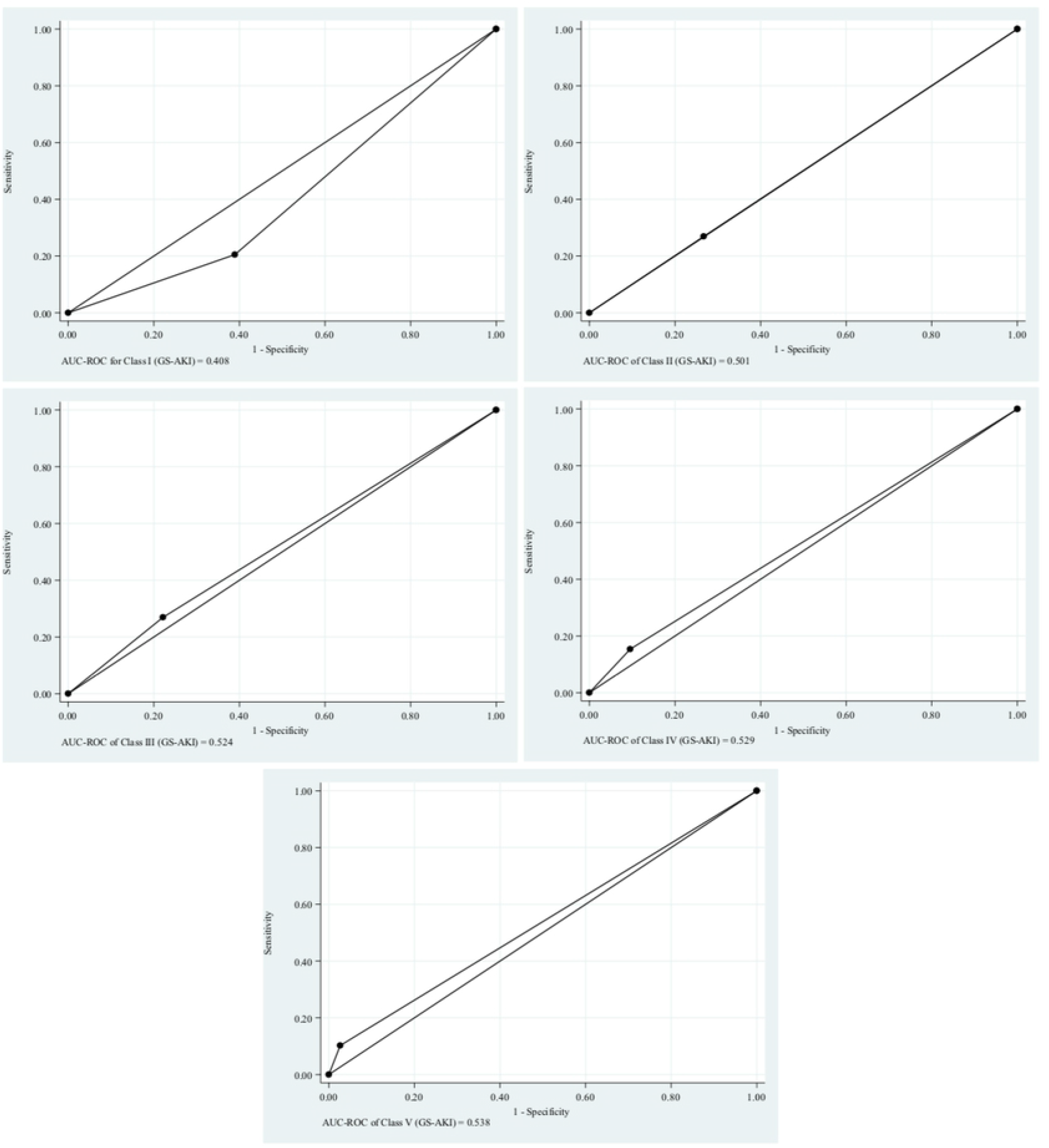
Area Under the Receiver Operating Characteristic (AUC-ROC) of the different SPARK Categories (N = 340)

## Discussion

Our study was performed using data on patients undergoing surgical procedures in a single center spanning three years. Based on the demographic data, the patient characteristics significantly associated with incidence of postoperative AKI include older age, pre-existing renal disease, longer duration of surgery, anemia, hypoalbuminemia, and hyponatremia.

Compared with SPARK index validation cohort, our study population had an older mean age and had more comorbidities, notably hypertension, diabetes, pre-existing renal disease, and had more patients using RAAS blocking agents, longer mean duration of surgery, more patients with preoperative anemia, dipstick albuminuria, hypoalbuminemia, hyponatremia, and higher incidence of AKI.

An external validation study for SPARK index (NARA-AKI cohort study) was done by Nishimoto et al.[17] in 2021 in a single center in Nara Medical University Hospital in Japan. Among the 5135 patients, 8.6% developed AKI. C statistic was 0.67 and 0.62 for AKI and critical AKI, respectively, which was similar to the results in our study. This population also had a higher mean age and presence of comorbidities compared to SPARK. Compared to this external validation, our study population had lower mean age. Our study also had higher incidence of hypertension and diabetes and had more patients with use of RAAS blockade, longer mean duration of surgery, anemia, hypoalbuminemia, dipstick albuminuria, and hyponatremia.

Compared with GS AKI, our study population had similar mean age, and proportion of hypertensive patients and with existing renal disease. Our study had higher rates of diabetes mellitus and congestive heart failure.

A local study was done by Doromal et al.[15] in 2016 in the University of Santo Tomas Hospital. The study included a total of 145 patients, with 59 patients who developed AKI. The c statistic was determined to be 0.799. Prevalence of post-operative AKI was 41%. They divided the risk stratification into seven groups as compared to the two groups in our study, which could be the reason they yielded a higher c statistic. Similar to our study, this study demonstrated that age and high baseline eGFR are be associated with significantly higher incidence of post-operative AKI. In Table 4, this study demonstrates that the higher the class for both SPARK and GS AKI, the higher the incidence of postoperative AKI.

To the best of our knowledge, this is the first study to compare the accuracy of SPARK and GS AKI on the same population. Our results are the following: SPARK had a sensitivity ranging from 17-43% and specificity ranging from 58-93% for Class B to C. GS AKI had a sensitivity ranging from 10-26% and specificity ranging from 61-97% for Class I to V. SPARK had a discriminative power (c statistic) ranging from 0.46 to 0.61 while GS AKI had a discriminative power ranging from 0.41 to 0.54. The c statistic is a simple method of determining if the model can accurately predict the outcome better than chance. Based on conventional cut-offs, the accuracy of both SPARK and GS AKI are fail. These are lower than the stated c statistics found in the original studies for both SPARK and GS AKI, which were stated to be 0.80, a good index. The difference could be due to several factors.

First, our data was collected from a single center with a limited sample size and number of AKI events. The prevalence of post-operative AKI was 22% in our study (N=340), 5.7% in SPARK cohort (N=90,805), 5.9% in NARA-AKI cohort (N=5135), and 1% in GS AKI cohort (N=56,519). Selection process is another factor that could have accounted for the differences of this study from the original SPARK and GS AKI studies. We excluded patients that did not fulfil the minimum data needed to determine the SPARK and GS AKI scores. Commonly unavailable data from our hospital database include serum albumin, urinalysis, and postoperative serum creatinine. These laboratories are not routinely requested for all surgical patients in this institution. This method of convenience sampling may have caused an overestimation of the incidence of postoperative AKI in our study population.

There are several limitations to this study. It was performed in a small sample of patients, with even fewer patients who could be eligible in the study due to the extensive laboratory workup required by SPARK. Convenience sampling was used because of the limited number of eligible patients.

## Conclusion

Based on our study, there is an association between higher risk classification in both SPARK and GS AKI and incidence of postoperative AKI. However, both clinical prediction models demonstrated poor measures of diagnostic accuracy such as sensitivity, specificity, PPV, and NPV, with a poor discriminative power to predict post-operative AKI in our study.

## Data Availability

All relevant data are within the manuscript and its Supporting Information files.

